# Hospital Resource Utilization in Epilepsy: Disparities in Rural vs Urban Care

**DOI:** 10.1101/19002766

**Authors:** Wyatt P. Bensken, Gina Norato, Omar I. Khan

## Abstract

**Objective:** This study assessed differences in resource utilization during epilepsy and seizure related hospitalizations in urban versus rural environments, to identify potential disparities in care for people living with epilepsy in rural communities.

**Methods:** A 10 year (2001 to 2011) state-wide hospital discharge database was used. Cost, length of stay, illness severity, and procedure use were compared between urban facilities, rural facilities, and epilepsy centers. Comparison across three separate years helped to identify differences between facility type as well as to assess practices changes within facilities over time.

**Results:** Average total charges between the three types of facilities was significantly different, with epilepsy centers having the highest average charge per patient, followed by urban and rural facilities (corrected p-values < 0.001). Illness severity was similar between the three groups, with the exception that epilepsy centers had a higher proportion of major severity cases. Length of stay remained fairly consistent across all three years for epilepsy centers (mean: 4.75 days), while decreasing for urban facilities (means: 4.39 to 3.72) and ultimately decreasing for rural facilities (mean: 3.31 to 3.09). Rates of procedure utilization revealed CT scan use persisted longer at rural facilities compared to urban facilities, while EEG and vEEG was low at rural facilities. vEEG was relatively restricted to epilepsy centers and had an initial substantial rise in use over the years and was associated with corresponding greater surgical procedure rates.

**Significance:** This study indicates a measurable difference between epilepsy centers, urban facilities, and rural facilities in cost and procedure utilization while caring for patients during epilepsy- and seizure-related hospitalizations. If these conclusions are valid, they support that persons living with epilepsy who live in rural communities and utilize these services receive care that is considered suboptimal when compared to urban centers or epilepsy centers.

## 1. Introduction

For persons living with epilepsy (PWE) access to appropriate care is important from time of initial seizure through diagnosis and seizure control. The need for well-trained neurologists and epileptologists is present throughout this process as recommendations for treatment (both pharmacologic and surgical) rely heavily on the ability to properly identify events, localize onset, and understand seizure and epilepsy etiology. Although diagnostic tools including magnetic resonance imaging (MRI) and electroencephalogram (EEG) are now readily available in most US hospitals and outpatient treatment settings, their implementation may vary. However, more advanced tools which increase precision, such as long-term video-EEG (vEEG) are generally limited to facilities with high levels of epilepsy care as well as options such as surgical evaluation and treatment. Further, as research evolves so does the utilization of these different tests in seizure diagnosis, such as the discontinuation of the computed tomography (CT) for seizure and epilepsy evaluation. How often these tests are utilized can provide an important measure of how epilepsy care is being dispensed for PWE.

Hospital care becomes an important piece of the continuity of care for PWE in cases of first seizure, breakthrough, and especially those who have refractory epilepsy with breakthrough seizures. Most often it is not the complexity of disease that determines where acute or emergent care is obtained, but rather factors such as convenience and availability, among others.^1^ In the case for PWE, the geographic location of any hospital or epilepsy center becomes a determinant in what resources are available to the patient. Previous work in understanding the presence of geographic-related disparities have indicated that those living farther from epilepsy centers and in more rural environments are potentially less likely to receive specialized care.^2–4^ Presumptively, varying levels of care can result in delayed diagnosis and potentially decreased effectiveness, causing worse care outcomes. To understand the potential presence of these disparities, it is important to examine differences in epilepsy and seizure care between epilepsy centers, other urban facilities, and rural facilities.

This study set out to study disparities in resource utilization between rural and urban facilities using cost, length of stay, illness severity, and procedures. One would assume that care provided at hospitals with a well-established epilepsy center would differ in quality from other hospitals and comparing this with rural facilities may have important implications for the treatment of PWE.

## 2. Methods

To examine the question of resource utilization for epilepsy and seizure hospitalizations in urban versus rural environments, a large state-wide hospital discharge database was used to assess differences in cost, length of stay, illness severity, and procedure use.

### 2.1. Data Source and Inclusion Criteria

Data from the Texas Department of State Health Services Health Care Information Collection was obtained for 2001, 2006, and 2011. This public use data file contains discharge data that is collected from most hospitals, with the exception of those that meet specific exclusion criteria, such as exceedingly low bed. Records contain facility information, patient demographics, and associated hospitalization information.^5^ Facility identifiers were matched to their address via a listing of facilities obtained from THCIC, and further assigned a Rural-Urban Commuting Area Codes to establish the designation of the facility.^6^

Inclusion criteria for the current analysis were patient over 5 years of age and a principle diagnosis code of either seizure or epilepsy, using the ICD-9 diagnosis codes of: 345.xx (xx = 0, 00, 01, 1, 10, 11, 2, 3, 4, 40, 41, 5, 50, 51, 6, 60, 61, 7, 70, 71, 8, 80, 81, 9, 90, 91), 779.0, and 780.xx (xx=3, 31, 32, 33, 39). Under these criteria, the cohort was narrowed from 8,534,551 to a final cohort of 37,859 records (Table 1).

**Table 1:** Summary of discharge records included in study

|  | Total Discharges |  |  |  |
| --- | --- | --- | --- | --- |
|  | Year |  |  | Total |
|  | 2001 | 2006 | 2011 |  |
| All Discharges | 2,679,784 | 2,917,188 | 2,937,579 | 8,534,551 |

| Total Discharges, Epilepsy & Seizure (> 5 y/o) by Facility |  |  |  |  |
| --- | --- | --- | --- | --- |
|  | 2001 | 2006 | 2011 | Total |
|  | 11,011 | 12,450 | 14,398 | 37,859 |
| Epilepsy Centers | 818 | 892 | 1,107 | 2,817 |
| Urban Facilities | 9,450 | 10,870* | 12,783 | 33,103 |
| Rural Facilities | 665 | 685 | 503 | 1,853 |
| Not matched | 78 | 2 | 5 | 85 |

\*One record removed due to aberrant length of stay

### 2.2. Definitions

A dichotomous rural and urban delineation was established via RUCA codes using previously employed methodology, which takes into account the commuting areas as well as population and density.^7^ Further, a sample of three epilepsy centers were extracted from the urban category to compare the variation in resource utilization. Epilepsy centers are commonly regarded as the gold standard of epilepsy treatment, and the three chosen facilities were all in urban areas, had the highest volume of epilepsy and seizure records in Texas, and were accredited as level 4 centers (the highest designation) by the National Association of Epilepsy Centers.

There were a few variables of interest for this analysis. Total charges were the sum of accommodation and ancillary charges, both covered and non-covered. Length of stay was an integer value with a minimum of 1, that is derived from the difference between end of care and start of care in days. Illness severity was a four-level categorical variable (minor, moderate, major, extreme) assigned from the All Patient Refined (APR) Diagnosis Related Group (DRG) Grouper.^5^ Procedure codes were ICD-9-CM codes associated with each record and matched to their descriptions using a file containing codes and matching descriptions available from the Centers for Medicare and Medicaid Services.^8^

### 2.3. Procedure Codes

To assess differences in specific resource utilization, a selection of epilepsy-related procedure codes were examined. These included Video/Radio EEG, MRI of brain and brainstem, EEG, CT scan, Other brain excision, Brain lobectomy, and the Wada test. A rate (procedures per 1,000 epilepsy/seizure discharges) was established for each procedure for comparison across facilities and years and was further stratified by illness severity.

### 2.4. Statistical Analysis

Data visualization and analysis was conducted via R. The cost and length of stay variables were evaluated using medians and means as well as interquartile ranges and confidence intervals, respectively. Further testing of the differences in cost for each year were done using using Kruskal-Wallis test due to the skewed nature of cost. The results for each center type were Bonferroni corrected for 3 comparisons: epilepsy center vs. urban, epilepsy center vs. rural, and urban vs. rural.

This study was exempted from Institutional Review Board review by the National Institutes of Health (NIH) Office of Human Subjects Research Protection (OHSRP).

## 3. Results

### 3.1. Total Charges

There was a significant effect of facility type on total charges for epilepsy/seizure cases: epilepsy centers were more costly than urban, which were more costly than rural facilities (Table 2). These differences were observed each year – 2001 (F=168.8, p < 0.001), 2006 (F=93.2, p < 0.001), 2011 (F=128.5, p < 0.001) – and all pairwise comparisons across years were significant (p < 0.001) after Bonferroni correction for 3 comparisons.

**Table 2:**
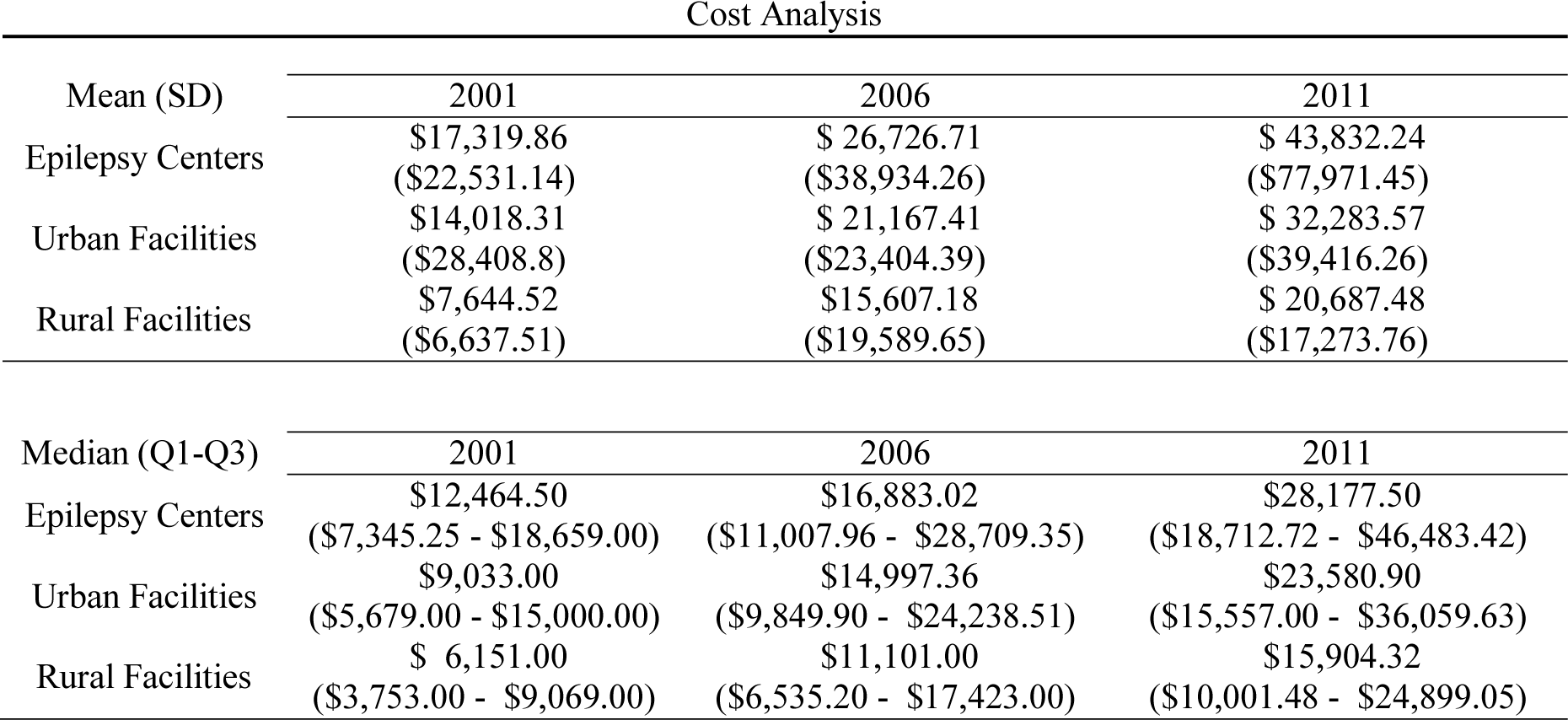
Cost analysis of total charges stratified by year and center

Epilepsy centers have seen a relative increase in their cost since 2001 that appears to continue through 2011, with urban facilities seeing a similar increase but a slower growth after 2006, and rural facilities cost leveling out with little growth in comparison (Figure 1). These trends seem to follow the cost trends for all discharges, however there does appear to be some slowing in rate of increase of mean charges for epilepsy in rural facilities when compared to their all discharge mean cost.

**Figure 1:**
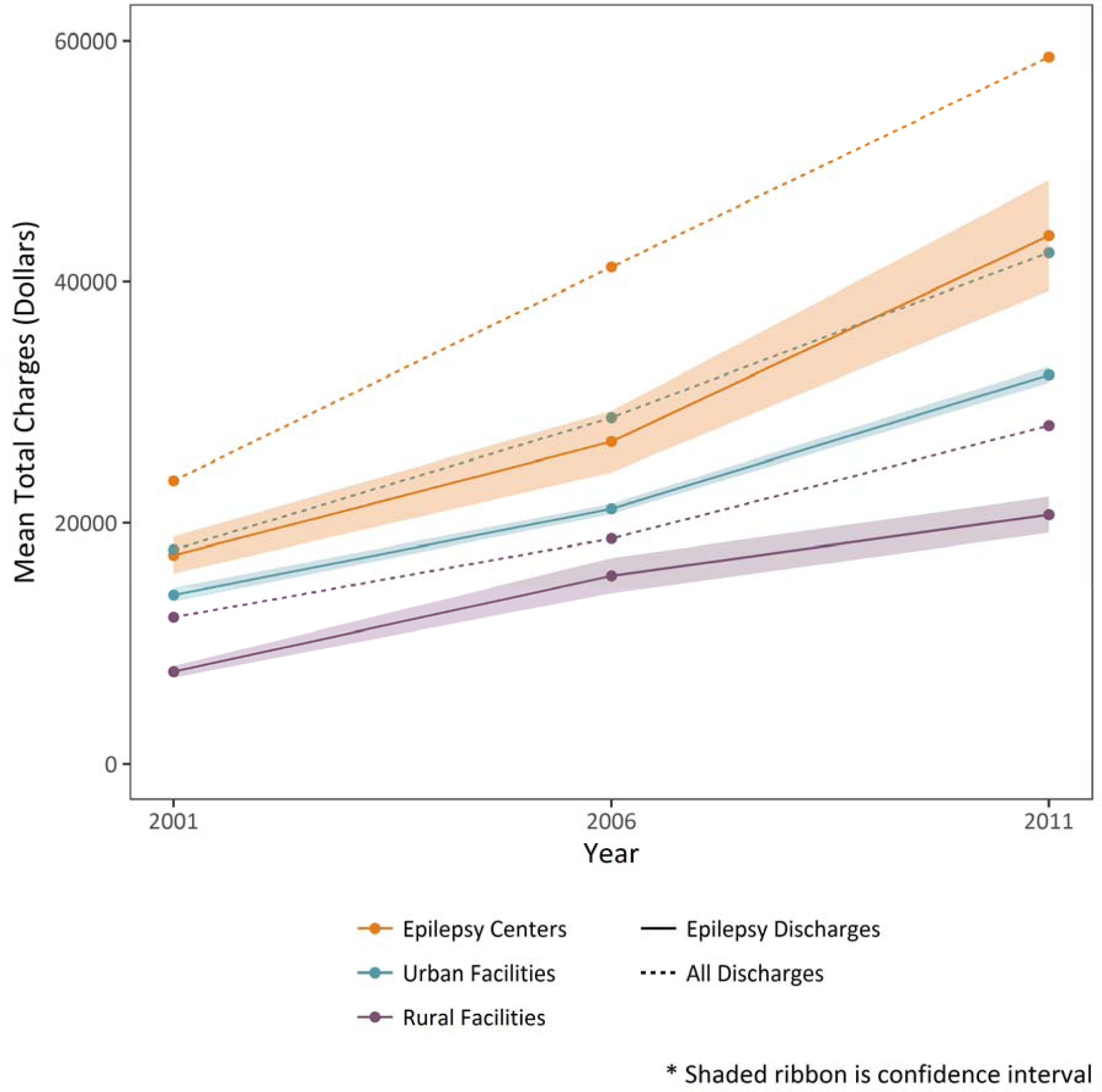
Mean total charges per discharge, with confidence interval, in US dollars. Solid lines represent epilepsy/seizure discharges >18 years old while dashed lines represent all discharges >18 years old

### 3.2. Illness Severity

Illness severity was comparable urban and rural facilities, while epilepsy centers had fewer moderate and more major cases – with the extreme and minor categories consistent across all three groups (Figure 2).

**Figure 2:**
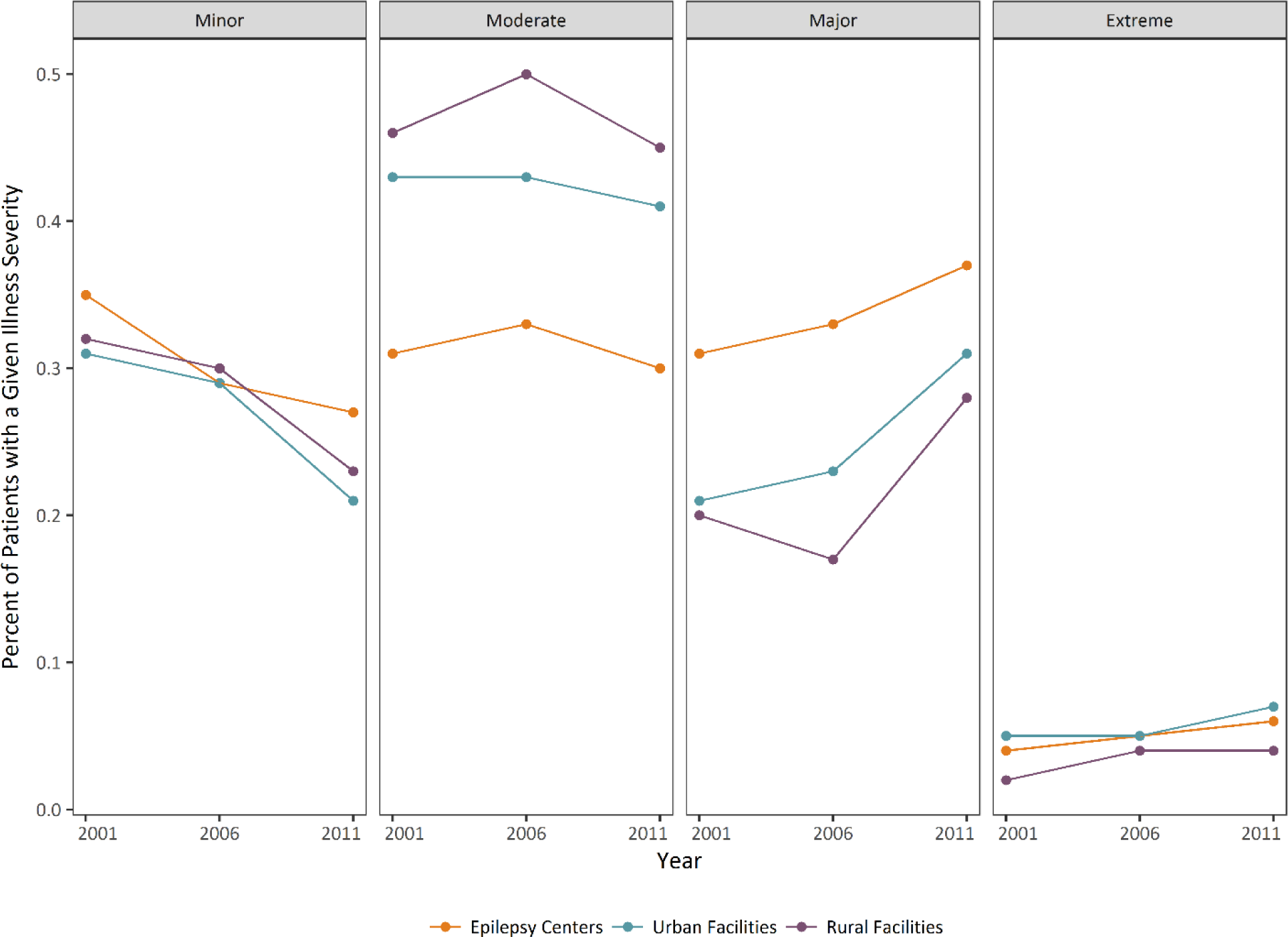
Trends of proportion of patients with a given illness severity, stratified by facility, across years

### 3.3. Length of Stay

The mean length of stay across the three years was consistent for epilepsy centers, only dropping from 4.73 (2001) to 4.70 (2011). Urban facilities decreased from 4.39 (2001) to 3.72 (2011). Rural facilities, on the other hand, saw an increase from 3.31 (2001) to 3.47 (2006) and then a decrease to 3.09 (2011). The medians were not as variable, with epilepsy centers having a median of 4 across 2001, 2006, and 2011 respectively, urban remaining at 3, and rural rising from 2 (2001) to 3 (2006) and then returning to 2 (2011).

### 3.4. Procedures

Rates of procedures indicate practice differences between and within facilities across time (Figure 3). Observed is the expected near-absent surgical procedure codes at urban and rural facilities, with changes in Wada testing at epilepsy centers, and a consistent rate of brain lobectomy over time. Further, CT scan as a diagnostic measure of epilepsy/seizure cases was observed to fall although this disuse was much slower at rural and urban centers than epilepsy centers. Finally, there is an observed increase in EEG at urban and rural facilities while vEEG experienced an increase then a relative leveling off at epilepsy centers. The rate of MRI use seemed to follow a trend of decrease use across all three facilities except for a stark increase from 2001 and 2006.

**Figure 3:**
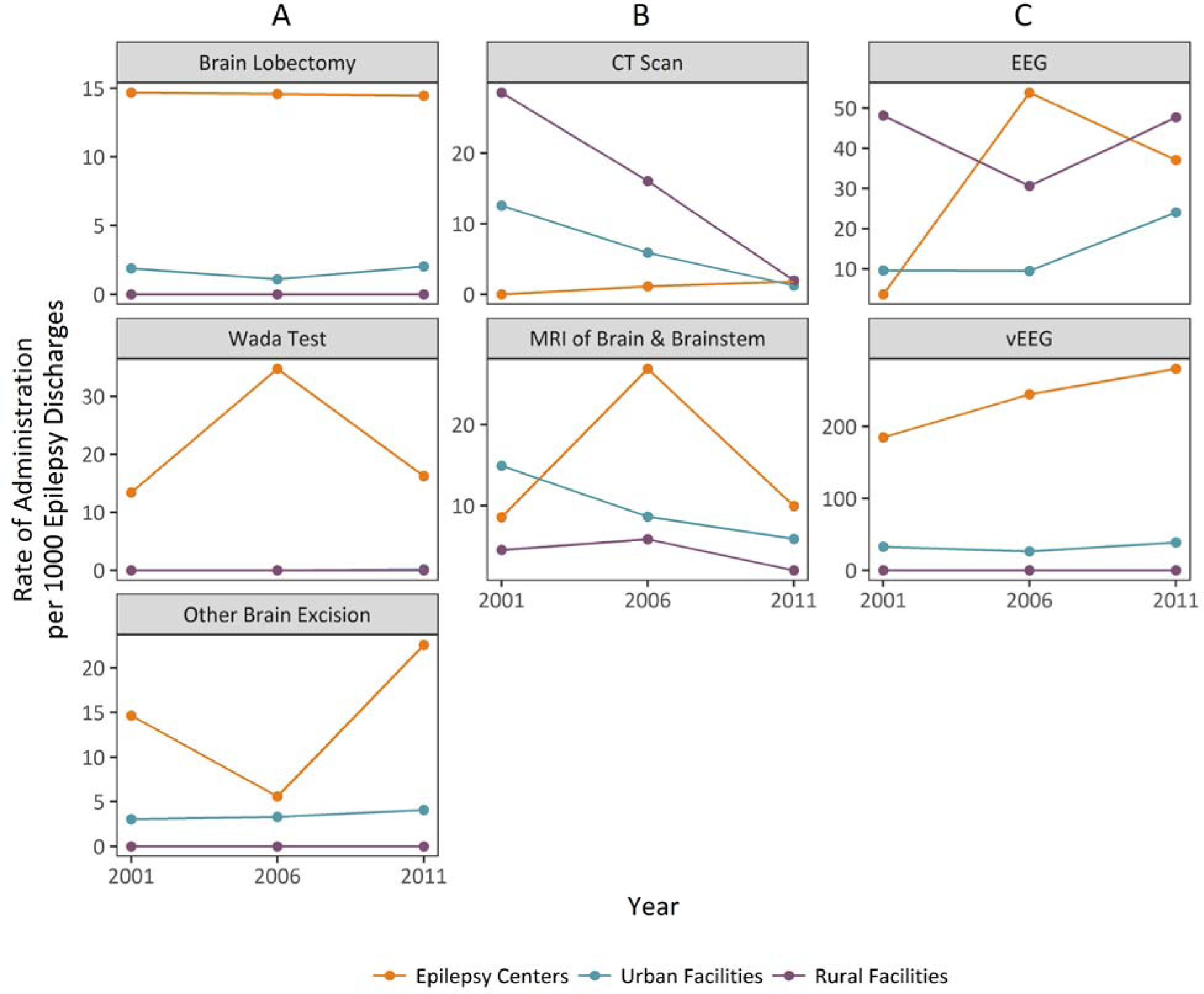
Rate of select procedures (per 1,000 epilepsy/seizure discharges). Column A is surgical procedures, Column B is diagnostic imaging, Column C is electrophysiological monitoring

### 3.5. Procedures by Illness Severity

Stratifying the rate of procedure codes by illness severity reveals key trends of resource utilization for the procedures that will provide the most insight on epilepsy care, specifically with EEG and vEEG (Figure 4). The rates of EEG at epilepsy centers increase and then decrease for minor, moderate, and major while the extreme severity saw a consistent increase. Contrarily, vEEG the epilepsy centers saw an increase in minor severity both over time and in comparison to urban and rural, while remaining consistent in extreme cases. The most notable differences for rural and urban facilities were in the use of the CT scan, where the highest rate was for moderate severity patients in rural facilities although that decreased to zero by 2011. Overall, these trends reveal that procedures are dependent on facility visited rather than a specific level of illness severity.

**Figure 4:**
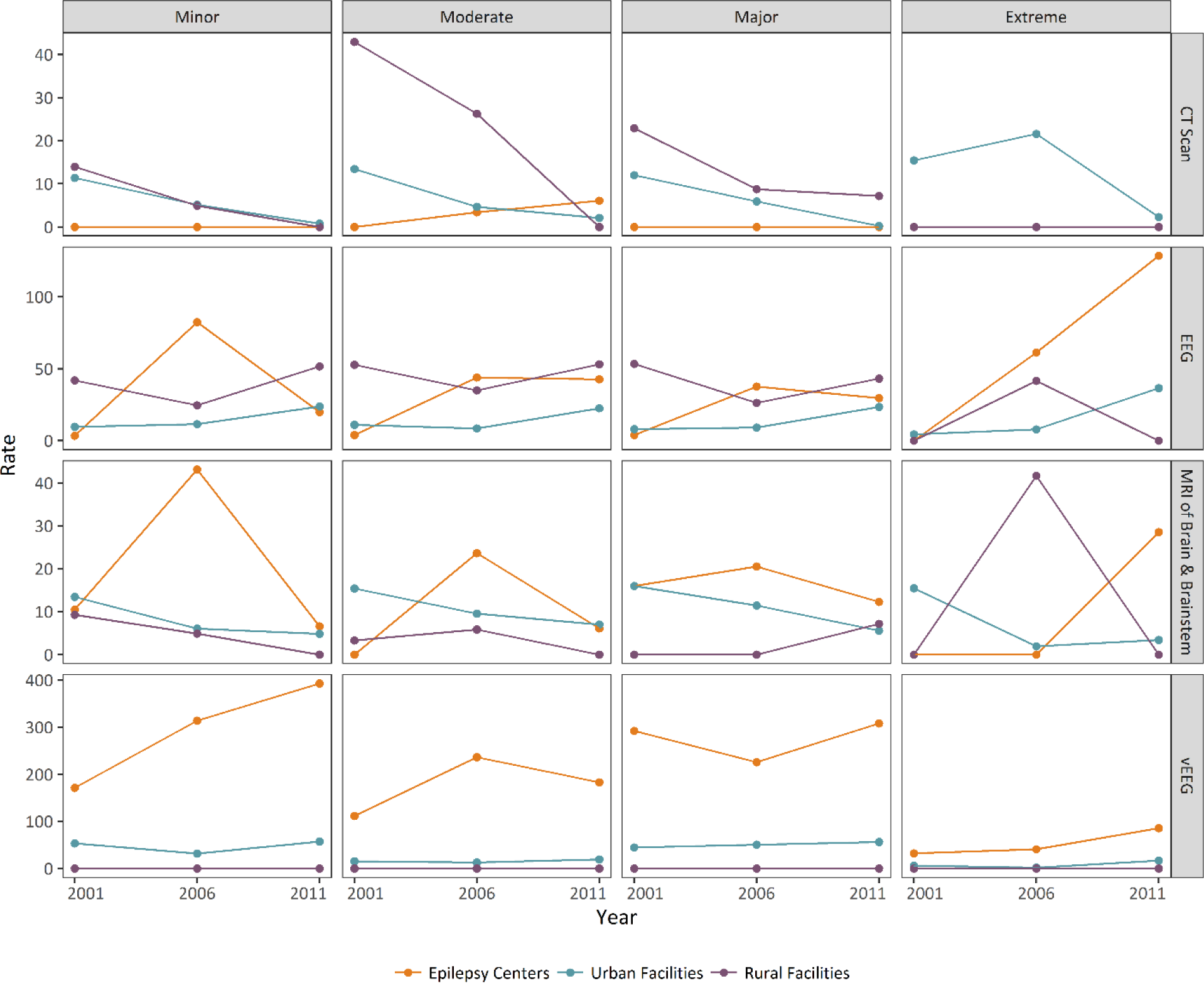
Rate of select procedures (per 1,000 epilepsy/seizure discharges) further stratified by illness severity

## 4. Discussion

Epilepsy can be a complex lifelong disease – treatment includes addressing control of seizures, medication costs and side effects, cognitive and psychiatric impairments as well as a social stigma that can have a significant impact on quality of life. Treatment can be particularly challenging when patients continue to have recurrent seizures and become refractory to standard anti-seizure drug regimens. Despite the wide variety and availability of diagnostic tools, as well as an increasing range of antiepileptic drugs and non-pharmacologic treatments, barriers still exists to provision of optimal comprehensive care.^4^

This study provides evidence for differences in resource utilization between epilepsy centers, urban facilities, and rural facilities. These differences are directly measured by cost between the facilities, as well as through length of stay, illness severity, and procedure use. These differences are most likely attributable to a lack of resources in rural environments, which should come as no surprise, and aligns with previous work in the field.^2,7,9,10^

The primary analysis of cost differences revealed that patients utilizing services in rural environments have a significantly lower cost of care compared to those utilizing services in urban environments and epilepsy centers (Figures 1 and 2). This difference in cost seems to be attributable not solely to differences in illness severity, but rather to differences in procedures that patients are receiving. Over time, these cost trends seem to follow the cost trends for the population as a whole and may present additional barriers for persons living with epilepsy such as restrictions on driving and work. Further, rural facilities have a lower standard deviation in cost per discharge than urban facilities, which is similarly lower than epilepsy centers. This difference is notable because it could perhaps indicate a limited variability and variety of care. With limited options and treatment, each case will be handled relatively the same and thus incur similar costs. However, at facilities with a myriad of diagnosis and treatment tools a high standard deviation would support a diversity in treatments. Whether this variation and use indicates an overuse at the epilepsy centers or an underuse at rural facilities is not clear, but regardless it provides evidence that differences do exist.

As expected, epilepsy surgery appears to be restricted to epilepsy centers and perhaps a small number of other urban facilities as shown by the absence of surgical procedures at rural facilities as well as a substantially lower rate at urban facilities when compared to epilepsy centers (Figure 3). However, using three time points, each five years apart, restricts our ability for a global view of time trends for epilepsy centers and the low counts contribute to high variability in the rates. Even so, the clear trends are that options for surgery are highly isolated to epilepsy centers and these centers did not appear to see any increase over time – perhaps further adding to challenges in accessing care. The use of the CT scans, and its high rate of use at rural facilities, underscores that rural facilities are using techniques that have been documented to be less accurate than other imaging alternatives for epilepsy investigation, although a decrease in MRI rates are observed in parallel.^11–13^ The two best measures of direct seizure and epilepsy care, EEG and vEEG, are clearly underutilized at rural facilities, further supporting limited care. Notably, the use of vEEG is limited to epilepsy centers, and a slow rise in urban facilities (Figure 4). Additionally, the use of vEEG was observed to be rising for minor illness severity cases – which is may include elective admissions for surgical evaluation. EEG use at epilepsy centers appears to be decreasing (Figure 3), however when stratified out there is a clear increase in extreme severity cases (Figure 4). These trends of EEG and vEEG use support that epilepsy centers are increasing their surgical evaluations and expanding their capabilities, and this is slowly spreading to other urban facilities, while rural facilities have been relatively stalled in their capabilities.

While this study was unable to directly measure the quality of care, length of stay analysis revealed that over time rural facilities have shown little improvement in length of stay trends with averages still around 3 days, despite evidence of no utilization of vEEG or other diagnostic or surgical procedures. This maintenance of length of stay indicates that over time direct care or indirect care (referral to other facilities) at rural facilities has not substantially changed.

Suboptimal care leading to health outcome disparities in rural areas is not a concept limited to epilepsy. This “mortality penalty” and its root causes, have been linked to rural environments since the 1980s, and work has shown rural residents having less access to emergency and preventative services, resulting in lower rates of resource utilization.^14,15^Unsurprisingly, there is also an increasing gap in life expectancy between residents of rural and urban communities, and highest death rates for children and young adults are for those in the most rural counties.^16^ In this context, the demonstrated differences in resource utilization for PWE further supports that for those living in rural environments access to ideal and appropriate care is limited.

The use of administrative data in epidemiological studies has limitations due to the fact that clinical information captured may not be as precise compared to data that is collected for research purposes.^17^ In this regard, and because of the reliability concerns of epilepsy and seizure diagnosis codes, it was not possible to guarantee that all records of patients included were true epilepsy cases. Including the seizure diagnosis code in this study ensured a broader group of patients was captured. This particular dataset has specific limitations in that facilities in the most rural areas are not captured in the required reporting requirements leading to potential underrepresentation of cases presenting in the most rural facilities. However, this limitation would likely mask the disparities and if these facilities were to be included we expect to see a larger disparity between epilepsy centers and urban facilities from the rural facilities. A similar issue with the facilities is that not all epilepsy centers or facilities with high level of epilepsy care were included in the epilepsy center sample group, and those facilities that were included were not necessarily epilepsy centers throughout the period studied. However, the epilepsy center group is meant as a gold standard comparison and in order to obtain a clean signal the three facilities selected were most likely to represent this ideal care setting. Despite these limitations, this study establishes a clear difference in resource utilization between epilepsy centers, urban, and rural facilities. Future studies should directly assess how these differences impact patient care outcomes, and more broadly what care is available at rural facilities for people living with epilepsy.

## Data Availability

Data used in this study is from the publicly available Texas Health Care Information Collection (THCIC).

https://www.dshs.texas.gov/thcic/

## 5. Acknowledgements

Funding: This research was supported by the Intramural Research Program of the National Institute of Neurological Disorders and Stroke (NINDS), National Institutes of Health (NIH).

## 8. Supplemental

**Supplemental Table 1:** Breakdown of illness severity by facilities and year

| Illness Severity, by Facility |  |  |  |  |
| --- | --- | --- | --- | --- |
| Epilepsy Center |  |  |  |  |
| Illness Severity | 2001 | 2006 | 2011 | Total |
| Minor | 286 | 255 | 303 | 844 |
| Moderate | 251 | 296 | 328 | 875 |
| Major | 250 | 292 | 406 | 948 |
| Extreme | 31 | 49 | 70 | 150 |
| Total | 818 | 892 | 1,107 |  |
| Urban Facilities |  |  |  |  |
| Illness Severity | 2001 | 2006 | 2011 | Total |
| Minor | 2,901 | 3,122 | 2,682 | 8,705 |
| Moderate | 4,093 | 4,702 | 5,274 | 14,069 |
| Major | 2,001 | 2,532 | 3,952 | 8,485 |
| Extreme | 453 | 510 | 875 | 1,838 |
| Total | 9,448 | 10,866 | 12,783 |  |
| Rural Facilities |  |  |  |  |
| Illness Severity | 2001 | 2006 | 2011 | Total |
| Minor | 215 | 203 | 116 | 534 |
| Moderate | 303 | 343 | 226 | 872 |
| Major | 131 | 114 | 139 | 384 |
| Extreme | 15 | 24 | 22 | 61 |
| Total | 664 | 684 | 503 |  |

\*Some discharges not included due to an unavailable illness severity

